# Which Pediatric Patients Still Experience Laboratory-Confirmed and Catheter-Related Bloodstream Infections: A Quality Analysis

**DOI:** 10.64898/2026.01.20.26344479

**Authors:** André Ricardo Araújo da Silva, Júlia Eimi Kitagima Tiba, Ludimila Frade Brandão Júlio da Silva

**Affiliations:** Materno Infantil-Department-Faculty of Medicine-Federal Fluminense University, Niterói, Brazil; Medicine Course-Federal Fluminense University, Niterói, Brazil

**Author notes:** Address for correspondence: Professor Gabizo Street 252, bloco 6, apt 501-ZIP-CODE: 20271-062, Maracanã-Rio de Janeiro.

**Keywords:** Children, Central line-associated bloodstream-infection (CLA-BSI), catheter

## Abstract

**Introduction:** Central line-associated bloodstream-infection (CLA-BSI) and catheter-related bloodstream infections (CR-BSI) remain a significant concern in pediatric inpatient units.

**Objective:** To analyze a case series of CLA-BSI and CR-BSI in hospitalized pediatric patients in hospitals with rigorous infection prevention measures.

**Materials and Methods:** This was an analytical, descriptive, and retrospective study conducted in patients aged 0 to 18 years, admitted between August 2023 and March 2025, with a diagnosis of CLA-BSI or CR-BSI in two pediatric hospitals in Rio de Janeiro, Brazil. Variables potentially associated with the occurrence of infection were analyzed.

**Results:** A total of 86 infections were evaluated, comprising 66 CLA-BSI and 20 CR-BSI. Sixty patients (69.8%) were male, with a mean age of 71.8 months. Sixty-five (83.7%) had previous comorbidities, 63 (73.2%) had a prior hospitalization, and 27 (31.4%) had another invasive device. The mean time from catheter insertion to infection diagnosis was 32.1 days, and the mean time from hospital admission to infection onset was 18.45 days. Gram-negative bacteria were isolated in 40/86 (46.5%) cases. At 30 days post-infection, 61/86 (70.9%) had been discharged, 20/86 (23.3%) remained hospitalized, and 5/86 (5.8%) had died. There was no correlation between the bacterial group and the type of catheter used (p=0.068), nor between infection type (CLA-BSI vs. CR-BSI) and mortality outcome (p=1).

**Conclusions:** CLA-BSI and CR-BSI occurred predominantly in patients with prolonged hospital stays and underlying comorbidities, and were mainly caused by Gram-negative bacteria.

## Introduction

The use of central venous catheters (CVCs) in hospitalized pediatric patients is both necessary and highly prevalent. These devices play an essential role in the diagnosis of diseases and serve as a widely employed therapeutic resource, particularly for medication administration and parenteral nutritional support. However, central venous catheters are frequently associated with infectious complications in the hospital setting.^1^ Consequently, Central line-associated bloodstream-infection (CLA-BSI) and catheter-related bloodstream infections (CR-BSI) are common in pediatric hospitalizations and represent an important cause of morbidity and mortality in this population.

CLA-BSI is a notifiable healthcare-associated infection (HAI) in Brazil and is the most frequent among hospitalized children using invasive devices. ^2,3^ Although CR-BSI is not subject to mandatory reporting, it remains associated with unfavorable health outcomes.^4^ From January 1, 2015, to December 31, 2020, a prospective, multicenter cohort surveillance study of HAIs across 630 intensive care units (ICUs) in 221 hospitals (including 68 pediatric facilities) from 123 cities in 45 countries reported that pooled mean rates of CVC-associated bloodstream infections varied significantly by ICU type. Pediatric ICUs demonstrated an incidence of 5.37 infections per 1,000 catheter-days, higher than the overall mean for all ICUs (4.55) as well as medical ICUs (4.62) and medical-surgical ICUs (4.66), but lower than trauma ICUs, which presented the highest incidence (11.94). Pediatric oncology ICUs presented an incidence of 4.37, similar to the global average, though lower than adult oncology ICUs (7.41), which ranked among the highest across clinical specialties. These findings suggest that, although pediatric patients experience above-average infection rates, relative risk is strongly influenced by the complexity of care and clinical profile, particularly in trauma and adult oncology settings.^5^ In Brazil, a 2015 study showed that among 889 patients diagnosed with at least one HAI episode, 341 died, corresponding to a mortality rate of 38.4%, underscoring the significance of HAIs as a public health concern. ^6^

In line with the national landscape, in 2024 the Brazilian Health Regulatory Agency (ANVISA) published Patient Safety and Quality in Healthcare Services Bulletin No. 32, presenting indicators of HAIs and antimicrobial resistance in Brazilian hospitals. The report highlighted 464 healthcare facilities that reported CLA-BSI in pediatric ICUs nationwide. The most frequently isolated microorganisms included coagulase-negative staphylococci, *Staphylococcus aureus*, and *Enterococcus faecium*, with resistance rates approaching 60% against major antimicrobials such as cephalosporins, carbapenems, ceftazidime, and polymyxins. These rates have shown a steady increase over recent years, emphasizing bloodstream infections as a critical and urgent healthcare issue.^7^

Several risk factors are associated with CLA-BSI and CR-BSI. In neonates, low birth weight and prematurity significantly increase the risk of catheter-related bloodstream infections.^8^ In the broader pediatric population, prolonged ICU length of stay, duration of catheterization, and comorbidities leading to immunosuppression are recognized contributors to infection. ^9,10^

To mitigate these risks, evidence-based care bundles have been implemented to prevent bloodstream infections. Recommended measures include avoiding femoral insertion sites, adhering strictly to hand hygiene protocols, correctly applying contact precautions, and removing unnecessary catheters.^11^ Despite the adoption of such bundles as a strategy for reducing CLA-BSI and CR-BSI, these infections still occur.^12^ Therefore, this study aimed to analyze a series of CLA-BSI and CR-BSI cases in pediatric inpatients hospitalized in facilities with stringent infection prevention practices. The results may inform more targeted interventions and support the development of strategies applicable even in high-risk patients, for whom bloodstream infections may still occur despite comprehensive preventive measures.

### Aim

To analyze a case series of CLA-BSI and CR-BSI in pediatric inpatients admitted to hospitals with rigorous infection prevention measures.

## Materials and Methods

This analytical, descriptive, retrospective study was conducted in two pediatric hospitals in Rio de Janeiro: ProntoBaby Children’s Hospital and Centro Pediátrico da Lagoa. Both are private institutions located in the city of Rio de Janeiro. ProntoBaby has 140 inpatient beds, including 45 intensive care beds (one unit dedicated to onco-hematology patients), and admits patients through its 24-hour emergency department as well as referrals from other institutions. Centro Pediátrico da Lagoa has 45 beds, including 20 intensive care beds, 16 surgical-cardiac intensive care beds, and the remaining beds in the general ward.

Patients aged 0–18 years admitted between August 2023 and March 2025 who met ANVISA diagnostic criteria (2025) for CLA-BSI or CR-BSI were included.^2^ Although CR-BSI is not a mandatory notifiable infection for ANVISA, both hospitals monitor it as a quality-of-care indicator. Exclusion criteria included transfer within 24 hours of diagnosis or unavailable outcome data.

Both hospitals have well-established Infection Control Committees (ICCs) since 2005 and adopted catheter-related infection prevention bundles in 2019, including mandatory bed rotation within 7 days. Additionally, both institutions have maintained antimicrobial stewardship programs since 2017, which include pre-authorization of restricted antimicrobials based on local flora and cost, daily multidisciplinary bedside discussions, infectious syndrome treatment guidelines, monitoring of antimicrobial consumption by clinical pharmacy, and antibiotic de-escalation guided by culture results.

The following variables were analyzed: sex, age, comorbidities, prior hospitalizations, presence of other invasive devices, site of infection, length of stay in the same bed, type of infected catheter, hospital length of stay, duration of catheterization, causative pathogen and resistance profile, and 30-day outcomes. Data were collected using Google Sheets and analyzed with descriptive statistics (frequencies, means, medians). Fisher’s exact test was applied to categorical variables and Mann–Whitney test to continuous variables. Statistical analysis was performed using Epi Info or similar software. A p-value <0.05 was considered statistically significant.

## Results

From August 2023 to April 2025, 85 bloodstream infections were diagnosed: 22 in 2023, 56 in 2024, and 7 in 2025. Of these, 65 were classified as CLA-BSI and 20 as CR-BSI. Sixty-eight infections (80%) occurred in ICU patients, while 17 (20%) occurred in ward patients. Among ICU cases, the incidence density of CLA-BSI was 54/16,777 (3.2 per 1,000 CVC-days), and CR-BSI incidence density was 8/16,777 (0.48 per 1,000 CVC-days). Table 1 summarizes patient demographics.

**Table 1.** Demographic data of patients with bloodstream infections in children and adolescents (Rio de Janeiro, 2023-2025)

| Variables | Results |
| --- | --- |
| **Age** |  |
| Mean in months (range) | 72.7 (0-210) |
| Median in months | 43.5 |
| **Sex** |  |
| Male – N (%) | 49 (57.6) |
| **Past Hospitalization Hystory N (%)** |  |
| Yes | 61 (71.8) |
| No | 21 (24.7) |
| Not reported | 3 (3.5) |
| **Comorbidity N (%)** |  |
| Yes | 66 (77.6) |
| No | 19 ( 22.4) |

Among 66 patients with comorbidities, 29/66 (43.9%) had hematologic malignancies or solid tumors, 13/66 (19.7%) had congenital heart disease, 11/66 (16.7%) had chronic neurological diseases, and 13/66 (19.7%) had other comorbidities. Table 2 presents data related to prior device use, length of stay, and quality indicators.

**Table 2.**
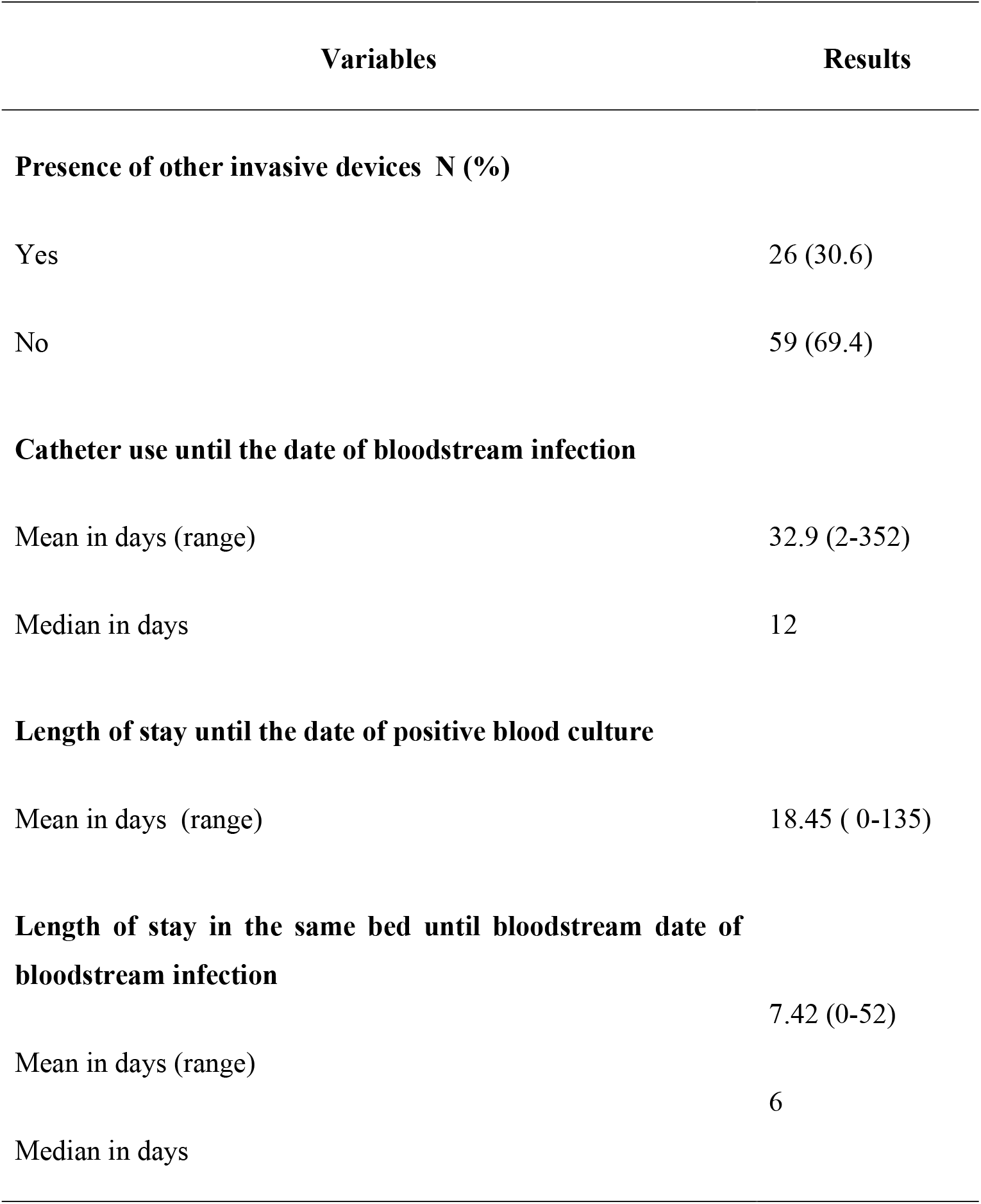
Use of Medical Devices Before and During Hospitalization and Associated Quality Indicators (Rio de Janeiro, 2023-2025)

Regarding catheter type, 63/85 (74.1%) infections occurred in patients with short-term CVCs, 18/85 (21.2%) in semi- or totally implantable central venous catheters, and 4/85 (4.7%) in peripherally inserted central catheters (PICCs).

Concerning causative agents, Gram-negative bacteria were isolated in 40/85 (47.1%), Gram-positive bacteria in 17/85 (20%), fungi in 13/85 (15.3%), mixed infections in 2/85 (2.4%), and no pathogen was identified in 13/85 (15.3%). Among the Gram-negative isolates, 26/40 (65%) demonstrated resistance to third- or fourth-generation cephalosporins or carbapenems, although only one case was caused by Pseudomonas aeruginosa. Resistance among Gram-positive isolates was 9/17 (52.9%).

At 30 days post-infection, 1/85 (71.8%) patients were discharged after infection, 19/85 (22.4%) remained hospitalized, and 5/85 (5.9%) had died. We also analyzed whether the presence of comorbidities could have influenced the outcome (death vs. survival). The results are presented in Table 3.

**Table 3.** Comorbidity in patients with bloodstream infection x survivals and not survivals (Rio de Janeiro, 2023-2025)

| Comorbidity in patients with bloodstream infection | Survivals<br>N=80 | Not survivals<br>N=5 | P value |
| --- | --- | --- | --- |
| Yes | 61 | 5 | 0.58 |
| No | 19 | 0 |  |

**Table 4.**
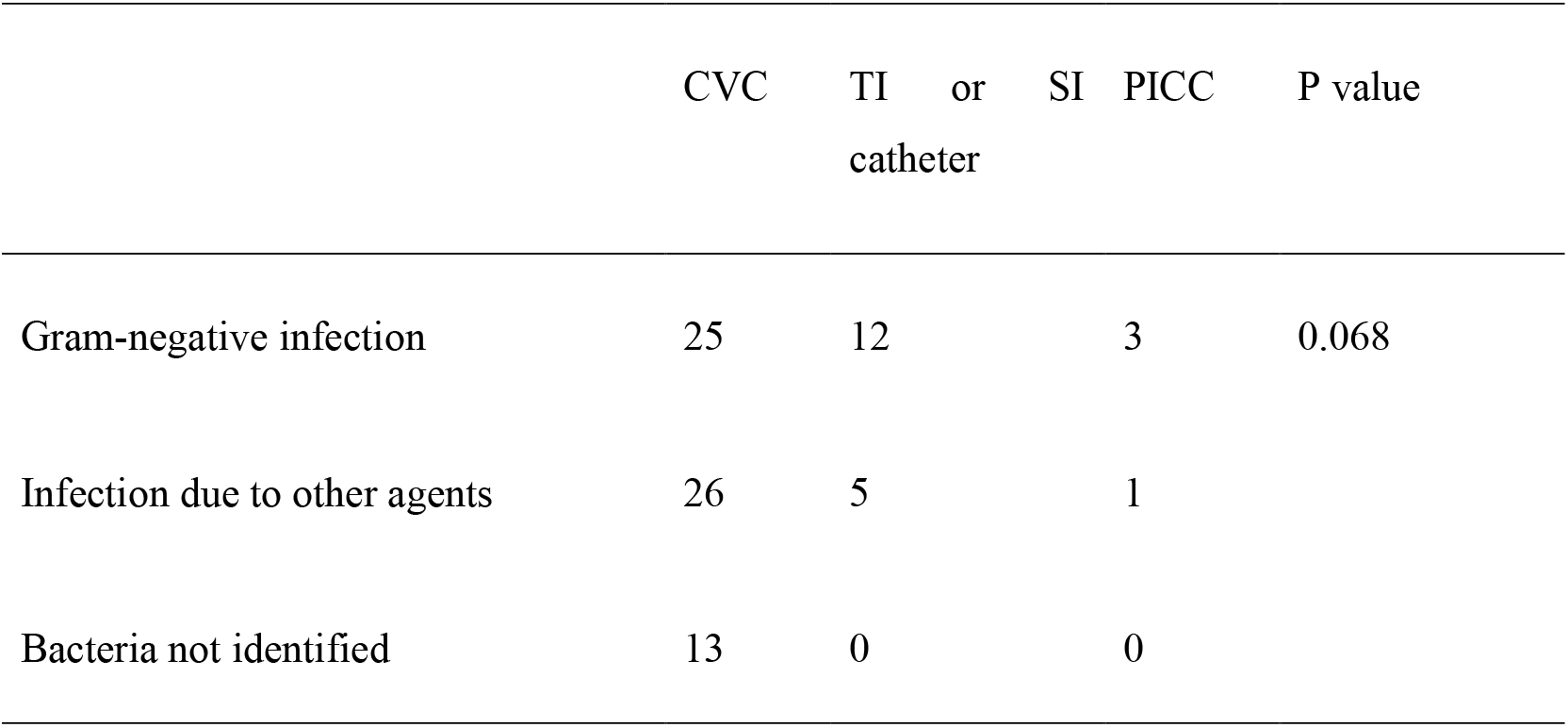
Influence of causative agent type on catheter type used (Rio de Janeiro, 2023-2025)

We also studied whether the type of comorbidity, specifically the most common groups identified (cardiopathies and neoplasms), had any influence on death or survival compared to other comorbidities. All 5 deaths occurred in patients with cardiopathies or neoplasms, while none of the patients with other comorbidities died (p = 0.1499).

No statistically significant difference was observed regarding deaths among patients with Central Line-associated bloodstream infections (CLA-BSI) (4/65, 6.2%) compared to those with catheter-related bloodstream infections (CRBSI) (1/20, 5%) (p = 1.00).

The following analysis addresses whether catheter infection by Gram-negative bacteria, other isolated pathogens, or cases in which no pathogen was isolated could have been influenced by the type of catheter used

At 30 days post-infection, 61/85 patients (71.8%) had been discharged, 19/85 (22.4%) remained hospitalized, and 5/85 (5.9%) had died. Analysis of comorbidities revealed that all deaths occurred in patients with congenital heart disease or malignancies, compared with none in patients with other comorbidities (p=0.1499). Mortality did not differ significantly between CLA-BSI (4/65, 6.2%) and CR-BSI (1/20, 5%) (p=1.00).

## Discussion

Central Line–Associated Bloodstream Infections (CLA-BSI) remain a significant challenge for healthcare systems, affecting both adult and pediatric inpatients, mainly due to their potential to increase length of hospital stay, healthcare costs, and mortality rates.^13^

Catheter-Related Bloodstream Infections (CR-BSI), although considered lower-impact events, highlight the need for continuous improvement in quality of care processes.^14^ For purposes of definition and nomenclature, infection surveillance systems in several countries, including Brazil, use CLA-BSI as the primary indicator for mandatory reporting.^2^

In this study, we investigated the occurrence of CLA-BSI and CR-BSI in a hospital that already adheres to rigorous infection prevention practices, including implementation and monitoring of central line insertion and maintenance bundles.

The adoption of multimodal measures—commonly referred to as bundles—has had a positive impact on reducing healthcare-associated infections, particularly CLA-BSI. A systematic review of studies published between 1990 and 2015 demonstrated a statistically significant reduction both in CLA-BSI incidence and central line utilization rates.^15^ Recommendations for catheter-related infection prevention are diverse and include standardized measures before, during, and after insertion.^16^ In our study setting, the institution has systematically implemented ANVISA’s recommended CLA-BSI prevention measures since 2019, with the exception of antibiotic-impregnated catheters.

Most detected cases occurred in patients admitted to pediatric intensive care units (PICUs), where central venous access is essential for the treatment of life-threatening conditions. ANVISA periodically publishes CLA-BSI incidence rates in Brazilian PICUs. However, it is important to note that many central lines inserted in PICUs remain in place after patients are transferred to general wards as their clinical condition improves, underscoring the need for continuous surveillance beyond the ICU.

When analyzing CLA-BSI cases that occurred in PICUs, the incidence density was 3.2 per 1,000 CVC-days—lower than the national rate for Brazilian PICUs in 2024 (4.1 per 1,000 CVC-days) and also below the multicenter global analysis by Rosenthal et al. (2024), which reported 5.37 per 1,000 CVC-days across 68 pediatric hospitals worldwide. ^5, 7^

The occurrence pattern of CLA-BSI and CR-BSI in our cohort was clearly defined: patients with underlying comorbidities, history of previous hospitalizations, and prior or concurrent use of invasive devices. These risk factors are consistently described in the literature. ^9,10, 17^ Such a hospitalization profile, characterized by high mortality risk and multiple comorbidities, is common in highly complex PICUs, where CLA-BSI incidence density is indeed higher when compared to children without comorbidities or with less complex clinical conditions. A similar finding was reported by Scarselli et al. (2021), who observed a CLA-BSI incidence density of 3.14 per 1,000 CVC-days among children with complex medical conditions, compared to zero among those without.^18^

When evaluating the pathogens causing CLA-BSI and CR-BSI, the local hospital flora must be considered. The most common causative agents are skin organisms, such as coagulase-negative *Staphylococcus* and *Staphylococcus aureus*, although Gram-negative bacteria are also frequently reported.^7^ In a Brazilian analysis of CLA-BSI in pediatric ICUs, the most frequent pathogens were *Klebsiella pneumoniae*, coagulase-negative *Staphylococcus*, and *S. aureus*, with a resistance rate of 37.28% among *K. pneumoniae* isolates.^7^ In our cohort, Gram-negative bacteria were the leading CLA-BSI pathogens, showing high resistance (65%) to third- or fourth-generation cephalosporins and carbapenems. Notably, only one case of carbapenem resistance was documented.

The study hospital admits a considerable number of patients with oncohematological diseases, those receiving home healthcare, and those with chronic neurological or pulmonary conditions. These patients usually require prolonged hospitalizations, which may have contributed to the observed bacterial resistance rates. Considering this hospital profile, we also investigated whether Gram-negative CLA-BSI occurrence was statistically associated with the type of catheter used; no such association was found, possibly due to the low number of infections in patients with PICCs. The use of PICCs has emerged as an interesting alternative for managing hospitalized patients across neonatal, pediatric, and adult populations, with studies demonstrating lower CLA-BSI rates compared to other central line devices. ^19,20^

Our study has several limitations. First, it reflects the reality of a single institution with well-established infection control and quality of care practices, where the patient profiles associated with CLA-BSI are already well defined. Second, it was not possible to analyze CLA-BSI incidence density among patients admitted to general wards, since mandatory surveillance is limited to ICUs. Nonetheless, our results remain valid and comparable with other institutions. Finally, as a descriptive and retrospective study, we did not evaluate risk factors among patients with central venous catheters who did not develop CLA-BSI or CR-BSI.

## Conclusion

CLA-BSI and CR-BSI occurred mainly in pediatric patients with prolonged hospital stays and comorbidities, predominantly caused by Gram-negative bacteria. In this high-risk population, CLA-BSI rates may not be completely eliminated despite strict preventive measures.

## Data Availability

All data produced in the present work are contained in the manuscript

## Ethical approval

The study was reviewd and approved by the Institutional Review Board (IRB), and was assigned the IRB log number CAEE 00678912.0.0000.5257. Throughout the course of this research, the core principles of respecting human anonymity and confidentiality, along with adhering to other ethical considerations, are upheld. The data extracted from the patients’ electronic medical records were de-identified, protected and kept carefully to ensure confidentiality.

## Declaration of conflicting interests

The author(s) declared no potential conflicts of interest with respect to the research, authorship, and/or publication of this article.

## Funding

The author(s) received no financial support for the research, authorship, and/or publication of this article.

## Notes

### Competing Interest Statement

The authors have declared no competing interest.

### Funding Statement

This study did not receive any funding

### Author Declarations

The study was reviewed and approved by the Ethics Committee of Hospital Universitário Clementino Fraga Filho (HUCFF/UFRJ), under number 00678912.0.0000.5257. Throughout the course of this research, the core principles of respecting human anonymity and confidentiality, along with adhering to other ethical considerations, are upheld. The data extracted from the patients' electronic medical records were de-identified, protected and kept carefully to ensure confidentiality.

